# Social Determinants of Health and Suicidal Behaviors among Children: U.S. Longitudinal Adolescent Brain Cognitive Development (ABCD) Study

**DOI:** 10.1101/2022.05.26.22275629

**Authors:** Yunyu Xiao, J. John Mann, Yu Hou, Julian Chun-Chung Chow, Timothy T. Brown, Paul Siu-Fai Yip, Alexander C. Tsai, Jyotishman Pathak, Fei Wang, Chang Su

## Abstract

**Importance:** Social determinants of health (SDoH) have well-characterized associations with child mental health outcomes. Their complex, multilevel influences on child mental health are less well understood.

**Objective:** To identify patterns across multiple domains of SDoH and estimate their associations with child mental health and suicidality outcomes over time.

**Design:** Panel study of 11 810 children aged 9 to 10 years who were enrolled in the Adolescent Brain Cognitive Development (ABCD) study and followed from September 1, 2016, and April 24, 2021.

**Setting:** Nationally-representative, population-based study across 21 sites in the U.S.

**Participants:** ABCD participants and their participating parents/guardians.

**Exposures:** 84 structural SDoH factors at baseline across 9 domains (discrimination, crime and drug use, education, health and environment, family type and disability, housing and transportation, minority status and language, socioeconomic status, and urbanization). We used unsupervised machine learning to identify patterns of clustering underlying the SDoH data.

**Main Outcomes and Measures:** Child mental health was measured with the parent-report Child Behavior Checklist. Suicidal ideation and suicide attempts were measured with child- and parent-report computerized versions of the Kiddie Schedule for Affective Disorders and Schizophrenia.

**Results:** Of 10 504 children included at baseline (median [SD] age, 9.9 [0.6] years), 5510 [52.5%] were boys and 4994 [47.5%] were girls; 229 (2.2%) were Asian, 1468 (14.0%) were Black, 2128 (20.3%) were Hispanic, 5565 (53.0%) were White, and 1108 (10.5%) were multiracial. Four SDoH patterns were identified: affluence (SDoH Pattern I); structural racism and discrimination (SDoH Pattern II); socioeconomic deprivation (SDoH Pattern III); and high crime, low education, and populated (SDoH Pattern IV) areas. Children with High Socioeconomic Deprivation at baseline (SDoH Pattern III) reported higher rates of externalizing (β, 1.43, 95% CI, 0.83, 2.02), internalizing (β, 0.75, 95% CI, 0.14, 1.37), and total (β, 1.16, 95% CI, 0.50, 1.81) problems, but these trajectories decreased over time. Children with High Structural Racism and Discrimination at baseline were the only group showing increasing trajectories of suicide attempts (OR, 1.42, 95% CI, [1.04, 1.93]) and depression (β, 0.19, 95% CI, 0.08, 0.29) over time. In contrast, living in affluent communities (SDoH Pattern I) was associated with lower rates of all internalizing and externalizing problems at baseline, but increasing trajectories of depression (β, 0.17, 95% CI, 0.09, 0.25), anxiety (β, 0.10, 95% CI, 0.02, 0.18), and withdrawal (β, 0.09, 95% CI, 0.01, 0.17) over time.

**Conclusions and Relevance:** Multiple domains of SDoH are associated with child mental health outcomes in cross section and over time. Targeted structural interventions may improve mental health outcomes and reduce suicide attempts among children.

**Key Points:** *Question:* What are the social determinants of mental health, suicidal ideation, and suicidal behavior among children in the U.S.?

*Findings:* In this cohort of 10 504 children, we used machine learning to identify four patterns of social determinants of health (SDoH). At baseline, socioeconomic deprivation was associated with internalizing and externalizing problems. Over follow-up, structural racism and discrimination were associated with suicide attempts.

*Meaning:* Multiple dimensions of structural interventions targeting different SDoH are needed to improve child mental health outcomes.

## Introduction

Social determinants of health (SDoH) have an increasingly recognized influence on the disparities in child mental health and suicidal ideation and suicide attempts (SI/SAs).^1–7^ Previous research has shown that county-level socioeconomic characteristics, such as poverty, unemployment, divorce rates, and social isolation, are associated with poorer mental health, greater rates of substance use, and higher suicide rates.^3,4,8–10^ In particular, higher county-level poverty was associated with greater suicide rates in children aged 5 to 19 years old, compared with counties with lower poverty rates.^6^ Rural areas had more firearm access and fewer mental health facilities and antidepressant medication prescriptions, which may contribute to rural-urban disparities in youth suicide.^5,7^ Understanding the associations between socioeconomic and structural factors and child mental health outcomes is critical to improving precision in designing social policies and programs.

SDoH is defined by the World Health Organization as the “conditions in which people are born, grow, live, work, and age”.^11,12^ Previous studies have tried to capture multiple SDoH by creating a composite index of contextual factors, including the Area Deprivation Index (ADI)^13^ and Social Vulnerability Index (SVI),^14^ and correlating these measures with other ecological measures of health (e.g., at the county or state levels).^3,4,8,9^ However, these index-based measures fail to capture the multidimensional nature of SDoH. Most indices only contain a limited number of variables (15 variables for ADI, 17 variables for SVI) and derive data from a single source (e.g., American Community Survey [ACS]). Driven principally by socioeconomic domains, few indices include measures that address structural racism (defined as the totality of ways in which society fosters discrimination through mutually reinforcing inequitable systems),^15^ which may be the fundamental cause of mental health disparities.^16–18^ In addition, few longitudinal studies existed to quantify the relationships between early childhood SDoH and subsequent trajectories of mental health.^19,20^ Such information would be critical to informing early detection effects to prevent mental and behavioral health problems.

This study aims to identify underlying patterns of SDoH among children using unsupervised machine learning techniques. Specifically, an SDoH pattern can be represented as a subgroup of children who have similar SDoH profiles. The premise of this manuscript is that a data-driven approach can identify meaningful SDoH patterns while addressing the heterogeneity and multicollinearity among multiple domains of SDoH. We further examine the associations between the SDoH patterns with subsequent mental health trajectories.

## Methods

### Study Design, Setting, and Population

Data were drawn from the Adolescent Brain Cognitive Development (ABCD) Study^21^. The baseline data are based on a stratified probability sample of schools from 21 metropolitan areas in the United States, from which 11 878 children were recruited in 2016-18. Children and parents completed surveys regarding their mental health, socioeconomic characteristics, and developmental outcomes. The marginal age, sex, and race/ethnic compositions for children across the data collection sites match closely with those of the U.S., as identified in the ACS from the US Census.^22^ More details about ABCD are described elsewhere.^21^

### Measures

#### Social Determinants of Health (SDoH)

We used ABCD linked external data, which includes geocoded population-level socioeconomic and environmental datasets derived from the primary residential addresses of the participating children. Drawing from multiple conceptual frameworks, previous empirical studies, and systematic reviews,^23–26^ we considered 84 contextual-level variables across five major areas defined by Healthy People 2030, including social and community context, economic stability, education access/quality, healthcare access/quality, and the neighborhood and built environment.^11^ We also reviewed existing SDoH composite indices (e.g., ADI,^27–29^ SVI,^14^ the Opportunity Atlas [OA],^30^ and the Child Opportunity Index [COI]^31^) and related literature^23–26^ to categorize our SDoH variables into nine domains, including bias (e.g., implicit and explicit attitudes towards race, sex, sexual orientation, immigrant status), crime and drugs (county-level counts of arrests and offenses from the Uniform Crime Reporting [UCR] dataset), education (e.g., educational and social resources, early childhood centers, educational level of area residents), health access and environment (e.g., access to healthy food, green space, walkability index as determined by the U.S. Environmental Protection Agency), household composition and disability (e.g., the proportion of persons over 65 years old, persons with disabilities, and other socioeconomically vulnerable groups), minority status and language (e.g., percentage of residents belonging to minority populations, percentage of residents with limited English proficiency), socioeconomic status (e.g., poverty rate, unemployment rate, percentage of residents without a high-school diploma), and housing type and transportation (e.g., percentage of mobile homes, crowded housing conditions). The categorization of SDoH follows the WHO SDoH framework and measurements of multidimensional structural racism.^11,12,32^ Detailed definitions and dimensions of SDoH can be found in **Figure 1** and **eTable 1** in the Supplemental Online Content.

**Figure 1.**
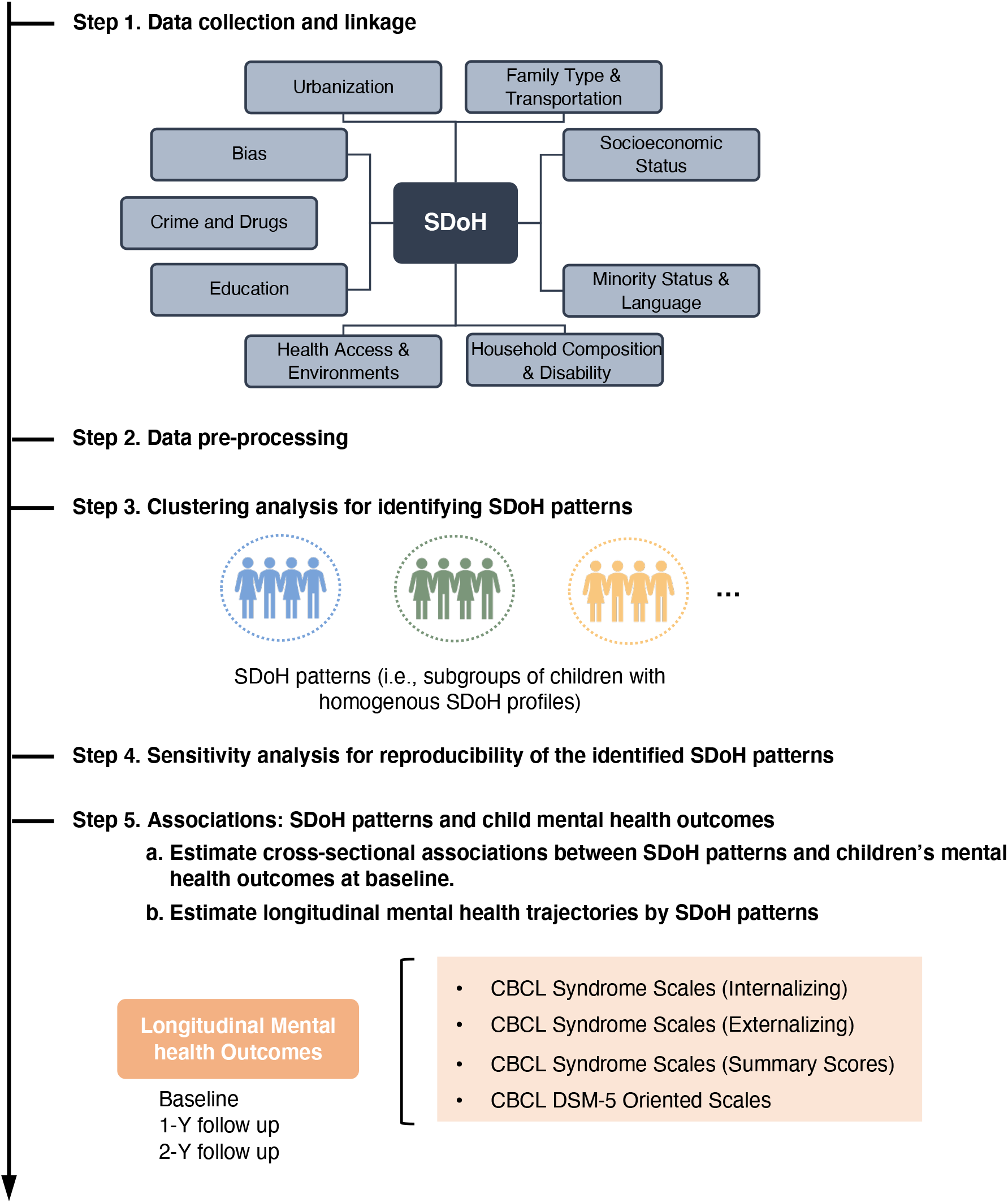
Study pipeline^a^. Abbreviations: SDoH, social determinants of health. CBCL, Child Behavior Checklist; DSM-5, Diagnostic and Statistical Manual of Mental Disorders, Fifth Edition. ^**a**^ We first collected 84 neighborhood-level SDoH variables from 9 domains for children in the ABCD cohort. After necessary data pre-processing, we performed clustering analysis to identify SDoH patterns. We performed sensitivity analyses to evaluate the reproducibility of the SDoH patterns. Finally, we estimated cross-sectional associations between SDoH patterns and children’s mental health at baseline and longitudinal mental health trajectories by SDoH patterns.

#### Child Mental Health Outcomes

We used the parent/caregiver-report Child Behavioral Checklist (CBCL),^33^ an empirically-validated and widely-used assessment, to measure children’s dimensional psychopathology and adaptive functioning. We included 8 individual scales for internalizing problems (e.g., anxious, withdrawn, somatic complaints), externalizing problems (e.g., rule-breaking, aggressive behaviors), and social-behavioral problems (e.g., thoughts, social, attention); 3 summary scores (internalizing, externalizing total problems); and 6 DSM-Oriented symptom subscales (depression, anxiety, attention deficits and hyperactivity, somatic problems, oppositional behavior, and conduct problems). Raw scores on this measure were converted to t scores using sex- and age-based norms derived from population-based studies. A high score reflects a more severe mental health problem, with a *T* score greater than 60 representing a clinically significant disorder.^34^ The rationale for using a parent/caregiver-report instrument for comprehensive mental health assessment has been discussed previously by the ABCD investigators.^33,35–37^ We used parent- and child-reported current and past suicidal ideation and suicide attempts from the Kiddie Schedule for Affective Disorders and Schizophrenia-Present and Lifetime Version using the *Diagnostic and Statistical Manual of Mental Disorders* (Fifth Edition) criteria (K-SADS-PL DSM-5).^38,39^ Any endorsement of the 18 suicidality-related items by either parent or child was included. All of the above mental health conditions were assessed at baseline and 1- and 2-year follow-up.

#### Sociodemographic Characteristics

Socio-demographic information about the participating children was based on parent/caregiver-reported sex, race/ethnicity (Asian, Black, Hispanic, Multi-Racial, white), parental marital status, household income, and family history of suicide and depressive disorders.

More details can be found in **Figure 1** and **eMethod 1** in the Supplemental Online Content.

### Statistical analysis

Among the 84 candidate SDoH variables, 82 are continuous variables and 2 are categorical variables. We assessed the missingness of all 84 variables and the distributions of the 82 continuous variables (see **eTable 1**). The 2 categorical variables were converted to binary variables (see **eTable 1**). Study participants with any missing values in the 84 variables were dropped from the analysis. The percentage of missingness ranged from 5-8% for nearly all variables. All continuous variables were scaled based on their z-score.

We assumed that a given pattern of SDoH can be represented by a subgroup of individuals (i.e., children in the ABCD cohort) who have homogenous neighborhood-level profiles of SDoH. To this end, we performed a data-driven cluster analysis to derive patterns of SDoH. After data preparation, hierarchical agglomerative clustering with Euclidean distance calculation and the Ward linkage criterion^40^ were applied to the SDoH variables. We used hierarchical agglomerative clustering because it is robust to different types of data distributions and typically produces a dendrogram that visualizes the data structure to help determine the optimal cluster number, and because it has shown promise in identifying underlying patterns from clinical profiles.^41–45^ Besides producing the dendrogram, we calculated 14 measures of clustering models provided by ‘NbClust’ software^46^ to determine the optimal number of clusters, i.e., patterns of SDoH. More details and rationale for these methods can be found in the **eMethods 2** and **3** in the Supplemental Online Content.

To evaluate the stability and robustness of the identified patterns of SDoH, we conducted several sensitivity analyses. First, we evaluated the extent to which our findings were sensitive to dropping participants due to missing values. This sensitivity analysis included children whose SDoH variables have a missing rate of < 70% to re-identify SDoH patterns. K-nearest neighbors (KNN) imputation^47^ was used to address missing values. Second, we evaluated sensitivity to the included data samples.

Specifically, we first split the entire cohort into 5 folds, then each time, we successively dropped 1 fold (20% children) and used the remaining 4 folds (80% children) to construct a subset.^48^ We then re-identified SDoH patterns in each subset. In both sensitivity analyses, patterns of SDoH were re-identified using the hierarchical clustering algorithm, following the same criteria as in the primary analysis for determining the optimal number of clusters. Finally, we compared the re-identified patterns of SDoH with those identified in the primary analysis (cluster labels from primary and sensitivity analyses were aligned based on a manual review of SDoH profiles of the clusters). We then assessed the extent to which there was overlap between the patterns of SDoH produced in the primary analysis vs. the patterns produced in the sensitivity analysis.

To interpret our identified patterns of SDoH, we visualized them in two ways: 1) Based on the 84 scaled SDoH variables, we calculated 2D representations for the individuals. We visualized them in 2D space using t-distributed stochastic neighbor embedding (t-SNE)^49^. We then visualized the identified SDoH patterns in the 2D t-SNE space (see **eMethod 4** in the Supplemental Online Content). 2) We used a chord diagram to show the SDoH characteristics of each identified pattern (see **eMethod 5** for details). We also characterized the identified SDoH patterns by evaluating their differences according to children’s demographics, family mental health histories, SDoH profiles, and prevalence of mental health conditions at baseline, and 1-year and 2-year follow-up. Data were presented as median (with interquartile range [IQR] for data that are not normally distributed) or mean (with standard deviation [SD] for data that are normally distributed) for continuous variables and exact patient number (with percentage) for categorical variables. To compare SDoH patterns, we performed analysis of variance (ANOVA) tests for continuous data and *χ*^2^ tests for categorical data. Two-tailed P-values smaller than 0.05 were considered as the threshold for statistical significance.

We estimated the associations between each of the identified baseline patterns of SDoH and the mental health conditions. For each mental health outcome with continuous values (e.g., the CBCL scales), we fitted a linear mixed effect model for each SDoH pattern, specifying member in the SDoH pattern as the explanatory variable of interest. For each outcome with binary values (i.e., suicide attempts or suicidal ideation), we fit a mixed effect logistic regression model separately for each SDoH pattern. For each of these regression models, study site (n=21) was included as a random effect, and baseline age, sex, and race/ethnicity were included as covariates.

We further evaluated changes in mental health outcomes (trajectories) by patterns of SDoH. For each pattern of SDoH, we fitted a linear mixed effects regression model (for the CBCL scales) or a mixed effects logistic regression model (for suicide attempts and suicidal ideation) to estimate trajectories for each mental health outcome. Age was specified as the explanatory variable of interest, with random effects for individuals and study sites. Sex and race/ethnicity were also included as covariates. Statistical significance was determined by a 2-sided P value <.05.

Data management and preprocessing were conducted in Pandas 1.2.1 (https://pandas.pydata.org) and NumPy 1.20.1 (https://numpy.org) in Python 3.7. Hierarchical clustering analysis and t-SNE were performed using SciKit-Learn 0.24 (https://scikit-learn.org/stable/) in Python 3.7 and NbClust in R 4.1. Chord diagrams were created using the R package ‘circlize’ (https://cran.r-project.org/web/packages/circlize/index.html). Mixed effects regression models were fitted using the R package ‘lme4’. All other statistical analyses were conducted using R version 4.1.

## Results

### Summary Characteristics of the Child Participants

The characteristics of the analytical sample are reported in Table 1. Of 10 504 children in the baseline sample included in this analysis (median [SD] age, 9.9 [0.6] years), 5510 [52.5%] were boys, 4994 [47.5%] were girls, 229 (2.2%) were Asian, 1468 (14.0%) were Black, 2128 (20.3%) were Hispanic, 5565 (53.0%) were white, and 1108 (10.5%) were multiracial children. Most children lived with married parents (7177, 68.3%). Nearly 26% (n = 2775) lived in families reporting an annual income of less than $50 000. More than half (52.5%, n = 5515) had a blood relative with a history of depression, and 567 (5.4%) had a parent who attempted or died by suicide.

**Table 1.** Baseline Characteristics of the Study Cohort and the Four Identified Social Determinants of Health Patterns.

| Characteristics | All | SDoH Patterns |  |  |  | P value<sup>c</sup> |
| --- | --- | --- | --- | --- | --- | --- |
|  |  | SDoH Pattern I:<br>Affluent<br>Communities | SDoH Pattern II:<br>High Structural<br>Racism and<br>Discrimination | SDoH Pattern III:<br>High<br>Socioeconomic<br>Deprivation | SDoH Pattern IV:<br>High Crime, Low<br>Education,<br>Populated |  |
|  | n = 10 504 | n = 4078 | n = 2661 | n = 2653 | n = 1112 | - |
| **Demographics** |  |  |  |  |  |  |
| Age, yr, mean (SD) | 9.9 (0.6) | 9.9 (0.6) | 9.9 (0.6) | 9.9 (0.6) | 9.9 (0.6) | <0.001 |
| Sex female, No. (%) | 4994 (47.5) | 1907 (46.8) | 1250 (47.0) | 1288 (48.5) | 549 (49.4) | 0.29 |
| Parents' education (< Bachelor), No (%) | 4176 (39.8) | 766 (18.8) | 981 (36.9) | 1754 (66.1) | 675 (60.7) | 0.01 |
| Race and ethnicity, No (%) |  |  |  |  |  |  |
| Asian | 229 (2.2) | 141 (3.5) | 15 (0.6) | 19 (0.7) | 54 (4.9) |  |
| Black | 1468 (14.0) | 178 (4.4) | 219 (8.2) | 999 (37.7) | 72 (6.5) |  |
| Hispanic | 2128 (20.3) | 449 (11.0) | 246 (9.2) | 760 (28.6) | 673 (60.5) | <0.001 |
| Multi-Racial | 1108 (10.5) | 416 (10.2) | 291 (10.9) | 297 (11.2) | 104 (9.4) |  |
| White | 5565 (53.0) | 2890 (70.9) | 1889 (71.0) | 577 (21.7) | 209 (18.8) |  |
| Parents' marital status, No (%) |  |  |  |  |  |  |
| Married | 7177 (68.3) | 3306 (81.1) | 2035 (76.5) | 1179 (44.4) | 657 (59.1) |  |
| Single parent | 2651 (25.2) | 647 (15.9) | 501 (18.8) | 1181 (44.5) | 322 (29.0) | <0.001 |
| Living with partner | 597 (5.7) | 119 (2.9) | 115 (4.3) | 251 (9.5) | 112 (10.1) |  |
| Household Income, No (%) |  |  |  |  |  |  |
| < \$50 000 per year | 2776 (26.4) | 382 (9.4) | 588 (22.1) | 1356 (51.1) | 450 (40.5) | |
| \$50 000 - \$100 000 per year | 2775 (26.4) | 978 (24.0) | 966 (36.3) | 606 (22.8) | 225 (20.2) | <0.001 |
| >\$100 000 per year | 4082 (38.9) | 2480 (60.8) | 969 (36.4) | 351 (13.2) | 282 (25.4) | |
| **Family history** |  |  |  |  |  |  |
| Blood relative has suicide history, No (%) | 2005 (19.1) | 754 (18.5) | 585 (22.0) | 502 (18.9) | 164 (14.7) | <0.001 |
| Blood relative has depression, No (%) | 5515 (52.5) | 2242 (55.0) | 1589 (59.7) | 1261 (47.5) | 423 (38.0) | <0.001 |
| Either parent with alcohol problem, No (%) | 1519 (14.5) | 546 (13.4) | 388 (14.6) | 429 (16.2) | 156 (14.0) | 0.012 |
| Either parent with drug use problem, No (%) | 3091 (29.4) | 1161 (28.5) | 890 (33.4) | 809 (30.5) | 231 (20.8) | <0.001 |
| Either parent with depression problem, No (%) | 1119 (10.7) | 301 (7.4) | 317 (11.9) | 391 (14.7) | 110 (9.9) | <0.001 |
| "Either parent been to a doctor or counselor<br>due to emotional/mental problem, No (%) | 4032 (38.4) | 1725 (42.3) | 1135 (42.7) | 892 (33.6) | 280 (25.2) | <0.001 |
| Either parent attempted or committed died by<br>suicide, No (%) | 567 (5.4) | 166 (4.1) | 174 (6.5) | 174 (6.6) | 53 (4.8) | <0.001 |
| **Mental Health Outcomes<sup>a</sup>** |  |  |  |  |  |  |
| CBCL Anxious/Depressed, mean (SD) | 53.46 (5.91) | 53.29 (5.69) | 53.95 (6.56) | 53.36 (5.72) | 53.13 (5.40) | <0.001 |
| CBCL Withdrawn/Depressed, mean (SD) | 53.47 (5.72) | 53.06 (5.20) | 53.62 (5.93) | 54.01 (6.24) | 53.30 (5.54) | <0.001 |
| CBCL Somatic Complaints, mean (SD) | 54.89 (6.04) | 54.42 (5.68) | 55.44 (6.37) | 55.18 (6.16) | 54.61 (6.05) | <0.001 |
| CBCL Social Problems, mean (SD) | 52.76 (4.68) | 52.26 (4.20) | 52.93 (4.91) | 53.40 (5.15) | 52.58 (4.37) | <0.001 |
| CBCL Thought Problems, mean (SD) | 53.77 (5.86) | 53.45 (5.48) | 54.39 (6.28) | 54.08 (6.27) | 52.76 (4.86) | <0.001 |
| CBCL Attention Problems, mean (SD) | 53.86 (6.11) | 53.41 (5.69) | 54.21 (6.43) | 54.45 (6.56) | 53.31 (5.51) | <0.001 |
| CBCL Rule-Breaking Behaviors, mean (SD) | 52.74 (4.83) | 52.11 (4.13) | 52.80 (4.77) | 53.79 (5.80) | 52.39 (4.38) | <0.001 |
| CBCL Aggressive Behaviors, mean (SD) | 52.79 (5.46) | 52.34 (4.87) | 53.05 (5.70) | 53.39 (6.16) | 52.39 (4.94) | <0.001 |
| CBCL Externalizing Problems, mean (SD) | 48.43 (10.59) | 47.78 (10.24) | 49.43 (10.78) | 48.70 (10.83) | 47.74 (10.58) | <0.001 |
| CBCL Internalizing Problems, mean (SD) | 45.68 (10.27) | 44.52 (9.77) | 46.43 (10.21) | 47.08 (10.96) | 44.80 (9.91) | <0.001 |
| CBCL Total Problems, mean (SD) | 45.81 (11.26) | 44.67 (10.75) | 46.98 (11.10) | 46.94 (11.95) | 44.52 (11.20) | <0.001 |
| CBCL DSM-5 Depression, mean (SD) | 53.58 (5.68) | 53.23 (5.28) | 53.97 (6.10) | 53.85 (5.94) | 53.26 (5.34) | <0.001 |
| CBCL DSM-5 Anxiety Disorders, mean (SD) | 53.46 (6.06) | 53.14 (5.61) | 53.96 (6.71) | 53.58 (6.18) | 53.12 (5.64) | <0.001 |
| CBCL DSM-5 ADHD, mean (SD) | 53.20 (5.60) | 52.74 (5.27) | 53.53 (5.78) | 53.78 (6.04) | 52.69 (5.04) | <0.001 |
| CBCL DSM-5 Somatic Problems, mean (SD) | 55.48 (6.61) | 55.01 (6.31) | 56.12 (6.93) | 55.73 (6.67) | 55.08 (6.57) | <0.001 |
| CBCL DSM-5 Opposition Behaviors, mean (SD) | 53.45 (5.36) | 53.15 (5.08) | 53.74 (5.56) | 53.77 (5.65) | 53.09 (5.11) | <0.001 |
| CBCL DSM-5 Conduct Problems, mean (SD) | 52.97 (5.44) | 52.30 (4.67) | 53.20 (5.60) | 54.04 (6.39) | 52.36 (4.65) | <0.001 |
| Suicidal Behaviors, No (%)<sup>b</sup> |  |  |  |  |  |  |
| Suicide Attempt | 161 (1.5) | 43 (1.1) | 42 (1.6) | 57 (2.1) | 19 (1.7) | 0.004 |
| Suicide Ideation | 1308 (12.5) | 515 (12.6) | 348 (13.1) | 318 (12.0) | 127 (11.4) | 0.45 |

Abbreviations: ADHD, Attention Deficit Hyperactivity Disorder; CBCL, Child Behavior Checklist; DSM-5, Diagnostic and Statistical Manual of Mental Disorders, Fifth Edition; SD, standard deviation; SDoH, social determinants of health.
<sup>a</sup> CBCL was used to assess children's internalizing problems (e.g., anxious, withdrawn, somatic complaints), externalizing problems (e.g., rule-breaking, aggressive behaviors), and three social-behavioral problems (e.g., thoughts, social, attention) according to parent or caregiver reports. We also included the CBCL DSM-Oriented symptom subscales for depression, anxiety, ADHD; somatic problems, opposition behaviors, and conduct problems. Raw scores on this measure were converted to t scores, with a t score greater than 60 representing a clinically significant disorder.
<sup>b</sup> Suicidal behaviors (suicide attempts and suicide ideation) were aggregation of parent- and child-reported outcomes.
<sup>c</sup> Comparisons across all 4 SDoH patterns were performed using analysis of variance (ANOVA) test for continuous variables and $\chi^2$ test for categorical variables. Two-tailed *P*-values smaller than 0.05 were considered as the threshold for statistical significance.

### Patterns of SDoH

We identified four patterns across the 84 indicators of SDoH (see **eResult 1** and **eFigures 1** and **2** in the Supplemental Online Content). To facilitate interpretation, greater levels of each domain and variables indicate poorer SDoH (visualized patterns in Figure 2 and summary statistics shown in Table 1). SDoH Pattern I (Affluent Communities, *n* = 4078, 38.8%) was characterized by higher socioeconomic status across all domains. SDoH Pattern II (High Structural Racism and Discrimination, *n* = 2661, 25.3%) was characterized by the highest level of bias and discrimination toward women, sexual minorities, and immigrants while having a larger proportion of mobile homes, group quarters (institutions and other group living arrangements, such as nursing homes and military barracks), and having a higher percentage of renters (rather than homeowners). SDoH Pattern III (High Socioeconomic Deprivation, *n* = 2653, 25.3%) was characterized by high rates of poverty, single mothers, people with disabilities, households without a vehicle, and traffic congestions in urban areas while having low access to healthy food and healthcare facilities. SDoH Pattern IV (High Crime, Low Education, Populated, *n* = 1112, 10.6%) had the highest rates of crime and drug sales, crowded housing, minority populations, and air pollution, while having low access to educational resources and poor quality education, green space, and homeownership. Results of sensitivity analyses confirmed the 4-cluster structure in the data. The patterns of SDoH that were reidentified in both sensitivity analyses overlapped highly with the patterns identified in the primary analysis. More details can be found **eResults 2, eFigures 3** and **4** in the Supplemental Online Content.

**Figure 2.**
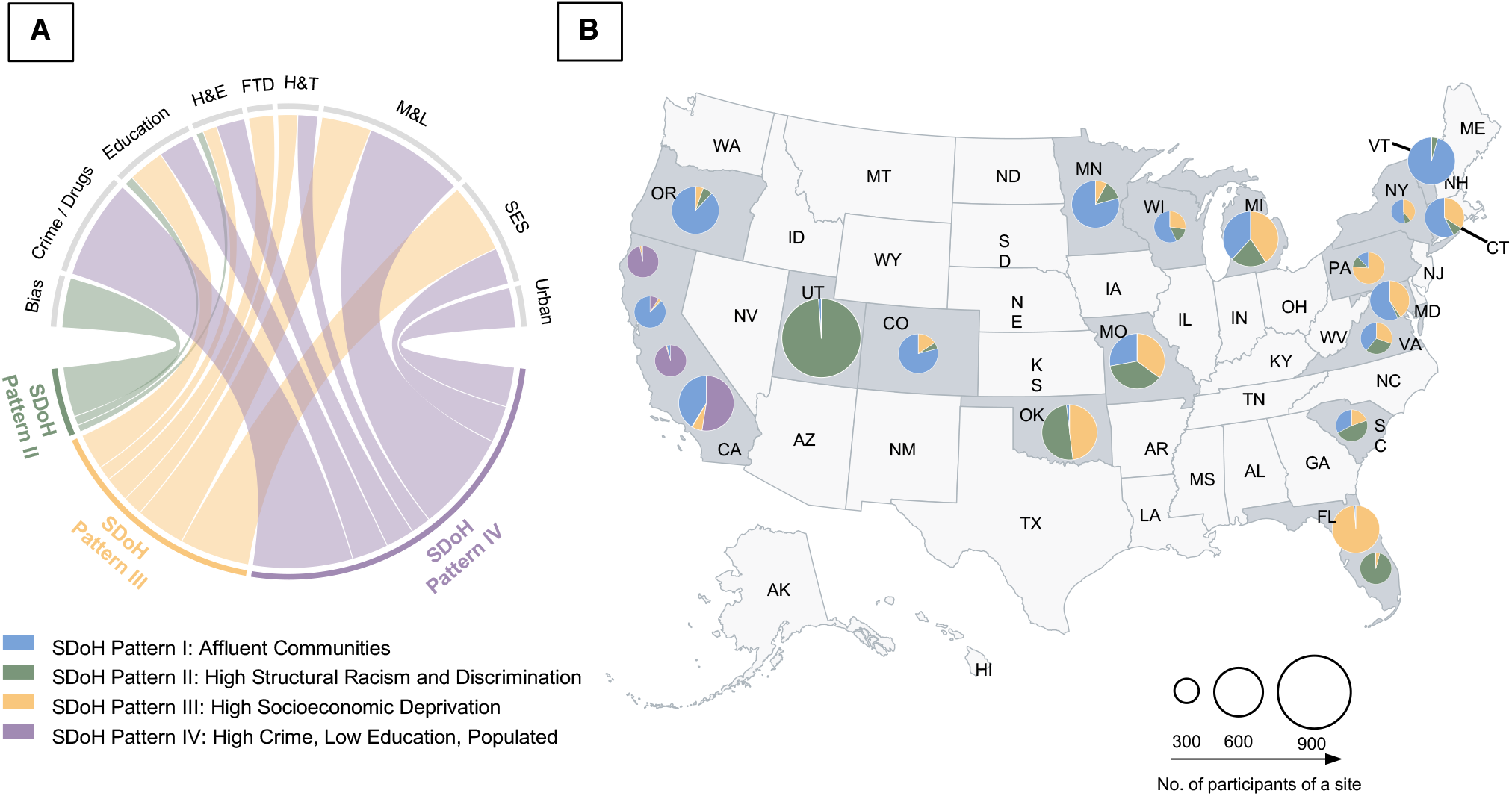
Characteristics of the identified social determinants of health (SDoH) patterns^a^. ^**a**^**A**. Chord diagrams showing differences in SDoH profiles across the 9 domains of the identified SDoH patterns. The width of a ribbon represents the proportion to the extent of disadvantaged status in a specific SDoH domain of a specific SDoH pattern. In other words, a broader ribbon between a SDoH pattern and a SDoH domain means the SDoH pattern has more disadvantaged status in the SDoH domain, compared to others. **B**. Geographic distributions of the SDoH patterns. Specifically, each pie indicates a specific site in the ABCD study, and the size of pie is proportional to the number of participants recruited from each site. Abbreviations: FTD, family type and disability; H&E, health and environment; H&T, housing and transportation; M&L, minority and language. SDoH, social determinants of health; SES, socioeconomic status.

### Associations between SDoH Patterns and Baseline Child Mental Health

Children living in SDoH Pattern III (High Socioeconomic Deprivation) reported significantly higher rates of baseline internalizing (β, 0.75, 95% CI, 0.14, 1.37), externalizing (β, 1.43, 95% CI, 0.83, 2.02) and total problems (β, 1.16, 95% CI, 0.50, 1.81), specifically for withdrawal/depression, somatic complaints, social problems, thought problems, rule-breaking problems, and problems of aggression (*P* <. 05, Figure 3). They were also more likely to have DSM-5 oriented depression (β, 0.36, 95% CI, 0.03, 0.68), somatic problems (β, 0.49, 95% CI, 0.11, 0.86), oppositional behaviors (β, 0.40, 95% CI, 0.10, 0.71) and conduct problems (β, 0.96, 95% CI, 0.64, 1.27). Children from SDoH Pattern II (High Structural Racism and Discrimination) also reported more somatic complaints (β, 0.41, 95% CI, 0.05, 0.77), thought problems (β, 0.36, 95% CI, 0.02, 0.70) and DSM-5 oriented somatic problems (β, 0.49, 95% CI, 0.11, 0.86), but did not have higher CBCL composite scores. Children who lived in affluent communities (SDoH Pattern I) had lower rates of all internalizing and externalizing problems, except for the anxious/depressed syndrome, and were the only group less likely to report suicide attempts (Odds Ratio [OR], -0.39, 95% CI, -0.76, -0.02).

**Figure 3.**
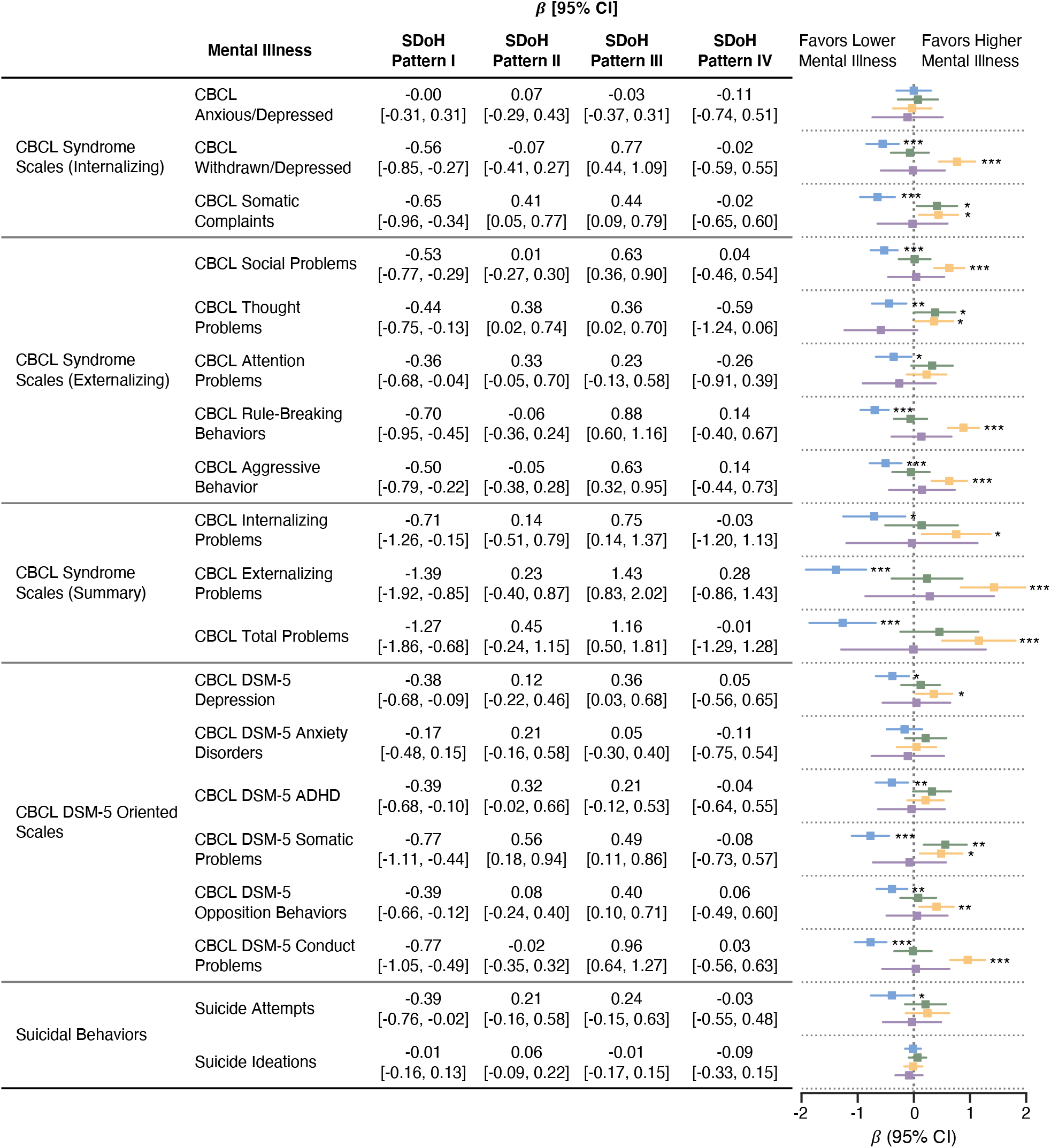
Forest plot showing associations between the identified social determinants of health (SDoH) patterns and mental health outcomes^a^. Abbreviations: ADHD, Attention Deficit Hyperactivity Disorder; CBCL, Child Behavior Checklist; DSM-5, Diagnostic and Statistical Manual of Mental Disorders, Fifth Edition. ^**a**^For outcomes in CBCL symptom scales (continuous), *β* (95% CI) and *P* values are based on linear mixed effect regressions analyses, adjusting for baseline age, sex, and race/ethnicity and including a random-effects term to account for within-site clustering; for suicidal behavior outcomes (binary), *β* (95% CI) and *P* values are based on was estimated based on mixed effect logistic regressions analyses, adjusting for baseline age, sex, and race/ethnicity and including a random-effects term to account for within-site clustering.

### Associations between SDoH Patterns and Child Mental Health Trajectories over Follow-Up

We found evidence of disparities in child mental health trajectories according to the different patterns of SDoH (Figure 4). Over time, children belonging to the SDoH Pattern II (High Structural Racism and Discrimination) group had increasing trajectories of DSM-5 oriented depression (β, 0.19, 95% CI, 0.08, 0.29) and the highest scores in internalizing problems, total problems, anxiety/depression, withdrawal/depression, somatic complaints, thought problems, attention problems, DSM-5 oriented anxiety disorders, ADHD, somatic problems, and suicidal behaviors. This group was the only group with an increasing trajectory of suicide attempts over time (OR, 1.42, 95% CI, [1.04, 1.93]).

**Figure 4.**
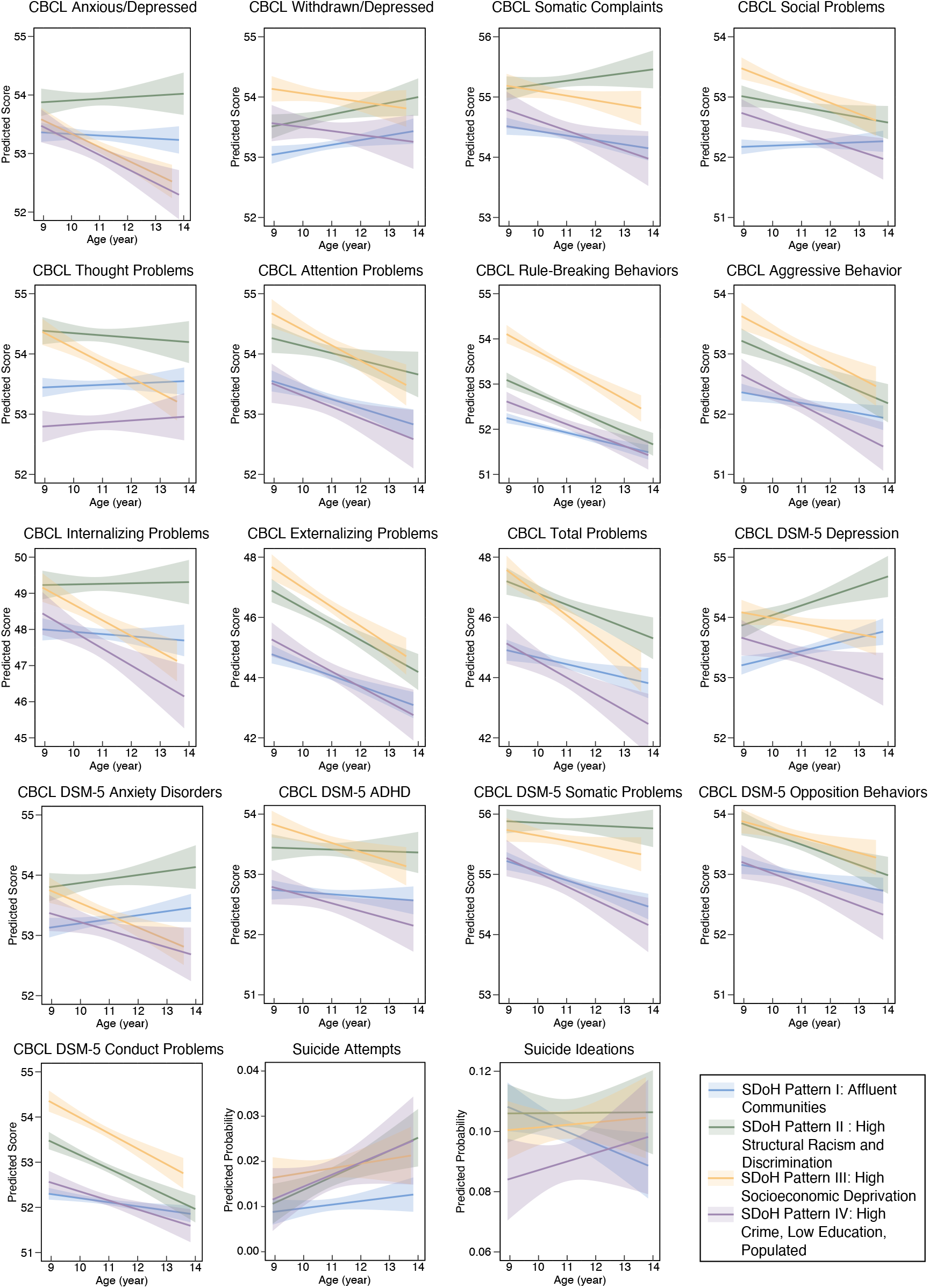
Estimated mental health trajectories by social determinants of health (SDoH) patterns^a^. Abbreviations: ADHD, Attention Deficit Hyperactivity Disorder; CBCL, Child Behavior Checklist; DSM-5, Diagnostic and Statistical Manual of Mental Disorders, Fifth Edition. ^**a**^For outcomes in CBCL symptom scales (continuous), *β* (95% CI) and *P* values are based on linear mixed effect regressions analyses, adjusting for baseline age, sex, and race/ethnicity and including a random-effects term to account for within-site clustering; for suicidal behavior outcomes (binary), *β* (95% CI) and *P* values are based on was estimated based on mixed effect logistic regressions analyses, adjusting for baseline age, sex, and race/ethnicity and including a random-effects term to account for within-site clustering.

Despite having lower rates of mental health problems at baseline, children living in affluent communities (SDoH I) had increasing trajectories of DSM-5 oriented depression (β, 0.17, 95% CI, 0.09, 0.25), DSM-5 oriented anxiety (β, 0.10, 95% CI, 0.02, 0.18), and withdrawal/depression (β, 0.09, 95% CI, 0.01, 0.17). On the other hand, their trajectories of suicidal ideation were decreasing over time (OR, 0.84, 95% CI, 0.77, 0.92). Children belonging to the SDoH III (High Socioeconomic Deprivation) group exhibited a contrasting pattern, with high levels of mental health problems at baseline but decreasing trajectories in all mental health measures, including internalizing problems (β, -0.52, 95% CI, -0.71, -0.34), externalizing problems (β, -0.57, 95% CI, - 0.75, -0.40), total problems (β, -0.75, 95% CI, -0.94, -0.57), and their subscales, as well as all DSM-5 oriented scales (e.g., anxiety, ADHD, somatic problems, oppositional behavior, and conduct problems, *P* < .05) except for DSM-5 oriented depression. Children belonging to the SDoH IV (High Crime, Low Education, Populated) group had increasing but nonsignificant trajectories of suicidal ideation over time.

## Discussion

Using data from a large, nationally representative panel study of children in the U.S., we used machine learning to derive 4 patterns of SDoH summarizing 84 different population-level socioeconomic and environmental variables. These patterns of SDoH were multidimensional, and differed in their levels of discrimination, crime and drugs, education, health and environment, family type and disability, housing and transportation, minority status and language, socioeconomic status, and urbanization. The clustering technique we used differs from other groupings methods, which have been restricted by a pre-defined threshold, have typically been restricted to a single domain of SDoH, or have relied on substantive knowledge about multiple domains.^50^ Importantly, our analysis revealed heterogeneity in the extent to which children are exposed to SDoH, as well as differential associations with mental health trajectories over follow-up. This information has important implications for screening, identification of child mental health problems, and intervention to prevent child mental health problems earlier in the life course. SDoH can be identified through publicly available datasets linked to the residential histories, and can potentially be useful in guiding early detection and treatment. Targeted social and public policies are needed to buffer children against the adverse effects of these social and structural influences on their mental health and development.^51^

Children living in **SDoH Pattern III (High Socioeconomic Deprivation)** show greater internalizing, externalizing, total problems, and their subscales at baseline. This is consistent with prior cross-sectional studies on the associations between deprivation and worse child mental health outcomes.^52–54^ Our findings strengthen existing knowledge by demonstrating the decreasing longitudinal trajectories between child mental health and area-level socioeconomic deprivation. During COVID-19, there were strong associations between area deprivation and greater stress among children.^55^ Early and comprehensive poverty alleviation programs can be promising tools to reduce the disparities in mental illness between children living in deprived areas and their better-off peers.^56^ Further investigations are needed to address the causal mechanisms linking socioeconomic deprivation and child mental health. It is important to evaluate the relative contributions of economic deprivations and individual family profiles (e.g., adverse childhood experiences, parental alcohol addictions, single-parent families). Additionally, children in SDoH Pattern III may also build up resilience in the long term, and exploring the protective factors against living in deprived areas is important to inform policy targets.^57^

Children living in **SDoH Pattern II (High Structural Racism and Discrimination)** had a notable risk of increasing suicide attempts trajectories, compared to the other SDoH Pattern. This is consistent with recent studies demonstrating the adverse impact of structural racism and perceived discrimination on child suicidal behaviors.^58,58^ **SDoH Pattern II** is also associated with greater trajectories of DSM-5 depression diagnosis. Our results are the first attempts to empirically support Krieger’s ecosocial theory of disease distribution in a child cohort, which describes how living in high structural racism and sexism contexts could embody mental illness towards minoritized populations in the life course.^59–61^ We found high-structural racism contexts as an important SDoH factor. Mental health providers should be aware of structural discrimination’s detrimental effect on childhood mental health.^55,62,62,63^ These results shed led to new insights into multi-level policies and clinical interventions to reduce bias, stigma, and racism at the contextual level to improve child mental health.

Nearly 40% of the child samples were in **SDoH Pattern I (Affluent Communities)**. While they reported lower proportions and risks of most mental illnesses, they were the only group exhibiting increasing trajectories of DSM-5-oriented depression, anxiety, and withdrawn/depression syndrome. The high prevalence of anxiety disorders was similar to previous reports conducted among affluent, urban child samples in Britain^64,65^ and Brazil.^66^ Our findings addressed special attention to depression and anxiety among children from affluent communities, which may be derived from their academic and interpersonal stressors.^67–69^ Importantly, during the COVID-19, the pressures on academic performance and pandemic-related stressors may be special risks for adverse mental illness among children, even if they live in affluent communities.^70^ Universal mental health screening, as recommended by the US Surgeon General,^71^ is important for all children to address the current mental health crisis, regardless of their families’ socioeconomic status.^72^ **SDoH Pattern IV (High Crime, Low Education, Populated)** did not show significant associations with child mental illness at the baseline. However, they reported increasing suicidal ideation trajectories. Recent community-based studies, mostly cross-sectional and among adults, have found emerging associations between area-level violent crime rates and suicide death rates.^73,74^ Our study is one of the first to demonstrate the longitudinal associations between high-crime exposure and suicidal ideation among pre-adolescent children.

### Strengths and Limitations

The strengths of our study include the use of data-driven techniques to identify underlying patterns of SDoH across wide-ranging, multidimensional indicators in a large, population-based, longitudinal sample of children. Our measures of child mental health outcomes are comprehensive and consistent with the DSM-5.^33,35^ However, interpretation of our findings is also subjective to several limitations. First, all mental health measures were based on self-report. We included parent-report assessments for CBCL, measured annually, to capture the mental health trajectories over time, but potential discrepancies between the reporting of the children and parents remain.^75^ Second, the ABCD sample is not nationally representative, although it covers 17 states across 21 study sites. Third, our data on the SDoH were limited by what is available in the ABCD-linked external data. This is not expected to be a significant limitation given that we included 9 different domains following influential theoretical frameworks^12,25,55,76^ and our previous empirical results based on the ABCD data.^55^ As more SDoH variables become available to investigators, further probing of the reproducibility of our findings is warranted. Fourth, missing data were common for some variables included in the clustering models. We used a complete-case analysis, but sensitivity analyses showed similar results when excluding only variables with a high degree of missingness.

## Conclusion

Our study provides an innovative way to understand the complex linkages between contextual socioeconomic/environmental variables and child mental health. Modeling the underlying patterns of SDoH – rather than using a single composite deprivation index – may better capture the complexity and spatial heterogeneity of SDoH, and also improve our understanding of their unique relationships with child mental health and suicidal ideation and behaviors. The identified SDoH can potentially contribute to screening efforts to improve the early detection and treatment of mental health problems. The study findings also suggest that policymakers can improve equity in mental health and service utilization through the use of targeted social and economic legislation and other policies,^51^ such as those targeting economic development, job security, food security, and education, which can potentially reduce rates of externalizing problems among children living in socioeconomically deprived areas.^77^ Yet children in affluent communities with access to economic and healthcare resources also have worsening trajectories of depression and anxiety. Our novel approach using machine learning algorism to identify patterns of SDoH provides actionable information that key stakeholders can use when designing and targeting interventions.

## Supporting information

Supplemental Online Content

## Data Availability

Data used in the preparation of this article were obtained from the Adolescent Brain Cognitive Development (ABCD) Study (https://abcdstudy.org), held in the NIMH Data Archive (NDA). This is a multisite, longitudinal study designed to recruit more than 10,000 children age 9-10 and follow them over 10 years into early adulthood. The ABCD Study is supported by the National Institutes of Health and additional federal partners under award numbers U01DA041048, U01DA050989, U01DA051016, U01DA041022, U01DA051018, U01DA051037, U01DA050987, U01DA041174, U01DA041106, U01DA041117, U01DA041028, U01DA041134, U01DA050988, U01DA051039, U01DA041156, U01DA041025, U01DA041120, U01DA051038, U01DA041148, U01DA041093, U01DA041089, U24DA041123, U24DA041147. A full list of supporters is available at https://abcdstudy.org/federal-partners.html. A listing of participating sites and a complete listing of the study investigators can be found at https://abcdstudy.org/consortium_members/. ABCD consortium investigators designed and implemented the study and/or provided data but did not necessarily participate in the analysis or writing of this report. This manuscript reflects the views of the authors and may not reflect the opinions or views of the NIH or ABCD consortium investigators.
The ABCD data repository grows and changes over time. The ABCD data used in this report came from ABCD Release 4.0 (DOI: 10.15154/1523041). DOIs can be found at http://dx.doi.org/10.15154/1523041.
Additional support for this work was made possible from supplements to U24DA041123 and U24DA041147, the National Science Foundation (NSF 2028680), and Children and Screens: Institute of Digital Media and Child Development Inc. The ABCD data used in this report came from the Adolescent Brain Cognitive Development Study (ABCD) Data Release: COVID Rapid Response Research (RRR) Survey First data release (Surveys #1, 2, and 3) (DOI: 10.15154/1520584) and COVID Rapid Response Research (RRR) Survey Second data release (DOI: 10.15154/1522601). DOIs can be found at https://nda.nih.gov/study.html?&id=1041 and http://dx.doi.org/10.15154/1522601

https://nda.nih.gov/abcd/

