## Supplemental Online Content for "Social Determinants of Health and Suicidal Behaviors among Children: U.S. Longitudinal Adolescent Brain Cognitive Development (ABCD) Study"

### eMethod 1 Definition of mental health outcomes

**Child Behavior Checklist (CBCL)[1] - Parent /Caregiver Report**
CBCL was used to measure children's dimensional psychopathology and adaptive functioning.
We included **8 individual scales:**
- Internalizing problems: anxious, withdrawn, somatic complaints;
- Externalizing problems: rule-breaking, aggressive behaviors;
- Social-behavioral problems: thoughts, social, attention.

We also included **3 summary scores:**
- Internalizing problems composite score;
- Externalizing problems composite score;
- Total problems composite score (summed scores for all 8 individual scales).

We also separately included **6 Diagnostic and Statistical Manual of Mental Disorders (DSM) -Oriented Scales** consistent with DSM diagnostic categories:
- Depression;
- Anxiety;
- ADHD;
- Somatic problems;
- Opposition behaviors;
- Conduct problems.

Raw scores on this measure were converted to **t scores** using sex- and age-based norms from population-based studies. A high score reflects a more severe mental health problem, with a t score greater than 60 representing a clinically significant disorder. CBCL is an empirically-validated and widely-used assessment. Detailed rationales of using CBCL for comprehensive mental health assessment from parents/caregivers were discussed previously[1-4].

**Suicidal behavior outcomes**
- **Suicide attempt** was determined any child- **OR** parent-reported suicide attempt.
- **Suicide ideation** was determined any child- **OR** parent-reported suicide ideation.

### eMethod 2 Agglomerative hierarchical clustering

In this study, agglomerative hierarchical clustering analysis was used to derive underlying SDoH patterns. On one hand, unlike other clustering methods like k-means clustering, agglomerative hierarchical clustering is usually robust as it's not sensitive to data distribution (e.g., k-means requires a sphere-like distribution of the data) and doesn't need an initialization procedure that may incorporate uncertainty. On the other hand, agglomerative hierarchical clustering typically produces a tree diagram known as dendrogram, which visually illustrates how the data points are agglomerated together in a hierarchical manner and distances between the clusters at different layers in the hierarchy, providing visible guidance in determining the optimal cluster number. For example, in **eFigure. 2**, each data point (i.e., child) is considered as a separate cluster at the beginning. Then, at each step, the two clusters that are most similar are joined into a single new cluster. The vertical axis of the dendrogram represents the distance or dissimilarity between clusters. High inter-cluster distance indicates a clear cluster structure of the data. Here, we performed agglomerative hierarchical clustering with Euclidean distance calculation based on clinical variable of the patients. Ward linkage criterion[5] was used to construct the hierarchy.

### eMethod 3 Determination of optimal cluster numbers for agglomerative hierarchical clustering

There is no golden standard for determination of optimal number of clusters in clustering analysis. To address this, we considered multiple criteria to determine the optimal cluster number in agglomerative hierarchical clustering analysis.

- Clusters were clearly separated in the dendrogram produced by agglomerative hierarchical clustering algorithm.
- Optimal suggested based on clustering measurements for the agglomerative hierarchical clustering model with Ward criterion, calculated by 'NbClust'[6], a famous R package for assisting a clustering method to determine the optimal cluster number of the data. In this study, we used 14 cluster measurements, including KL index[7], CH index[8], Hartigan index[9], Cindex[10], DB index[10,11], Ratkowsky index[12], Ball index[13], Ptbiserial index[14], Gap index[15], Frey index[16], McClain index[17], Dunn index[18], SDindex[19], and SDbw index[20].
- Cluster were clearly separated in low dimensional space.
- No cluster will dominant the studied cohort (containing over 50% individuals).

### eMethod 4 SDoH pattern visualization based on t-SNE plot

t-SNE (i.e., t-distributed stochastic neighbor embedding)[21] is a novel dimension reduction technique that are widely used for data visualization. Via a non-linear transformation, t-SNE is able to project the high-dimensional data into a low-dimensional space, such that close data points in the original space will be close in the low-dimensional space, and vice versa. We performed t-SNE on the 84 SDoH variables to visualize children in the 2-D space. Children's SDoH pattern memberships were colored in the t-SNE plots. t-SNE plots were created by the 'Scikit-Learn 0.24' and 'matplotlib 3.0' packages in Python.

### eMethod 5 SDoH pattern visualization based on the chord diagram

The chord diagram plot was used to visualize patterns of disadvantaged SDoH status of the identified SDoH patterns. The chord diagram was generated based on the 'Circlize' package[22] in R.

Specifically, we first grouped the SDoH variables into 9 SDoH groups. For variables like ECE enrollment of which lower values indicate the more disadvantaged status, we multiply the values with -1 to make sure higher values indicate the more disadvantaged status. Then, for each SDoH pattern, if median of a SDoH variable is higher than 75% percentile of the whole population, we added a ribbon with one unit width between the SDoH pattern and the specific variable group, normalized by variable number of the group. In this way, a broader ribbon between a SDoH pattern and a variable group means the SDoH pattern has more disadvantaged status in the SDoH group, compared to others. The 9 SDoH groups were listed as below.

- **Bias,** including state level indicators of sexism from survey and implicit bias measures, state level indicators of racism from survey and implicit bias measures and state level structural variables, state level indicators of bias against sexual orientation from structural variables, and state level indicators of immigrant bias from survey and implicit bias measures and state level structural variables.
- **Crime / Drugs,** including Residential history derived - Uniform Crime Reports: total adult offense, adult violent crimes, drug abuse violations total, drug sale total, Marijuana sale, drug possession total, and DUI.
- **Education,** including ECE Centers, High-quality ECE Centers, ECE enrollment, Third grade reading proficiency, Third grade math proficiency, High school graduation rate, Advanced Placement (AP) course enrollment, College enrollment in nearby institutions, School poverty, Teacher experience, and Adult educational attainment.
- **Health & Environment (H&E),** including Access to healthy food, Access to green space, Walkability, Housing vacancy rate, Hazardous waste dump sites, Industrial pollutants in air, water or soil, Airborne microparticles, Ozone concentration, Extreme heat exposure, Health insurance coverage, Estimated lead risk in census tract of primary residential address (1-10 scale), Spatio-temporal model predictions measured in μg/m3 at 1 km2 resolution, Spatio-temporal model predictions measured in ppb (parts per billion) at 1 km2 resolution, Spatio-temporal model predictions measured in ppb (parts per billion) at 1 km2 resolution.
- **Family type & Disability (FTD),** including Percentage of persons at least 65 years old, Percentage of persons 17 years old and younger, Percentage of civilian noninstitutionalized population older than age 5 with a disability, Percentage of single-parent households with children less than 18 years old.
- **Housing type & Transportation (H&T),** including Percentage of housing in structures with 10+ units, Percentage of mobile homes, Crowding, Percentage of households without a vehicle, Percentage of persons living in group quarters.
- **Minority status & Language (M&L),** including Percentage minority population (i.e., all but white, non-Hispanic) and Percentage of persons at least 5 years old who speak English "less than well".
- **Socioeconomic status (SES),** including Percentage of population aged >=25 years with <9 years of education, Percentage of population aged >=25 years with at least a high school diploma, Percentage of employed persons aged >=16 years in white collar , Median family income, Income disparity defined by Singh (2003), Median home value, Median gross rent, Median monthly mortgage, Percentage of owner, Percentage of occupied housing units with >1 person per room (crowding), Percentage of civilian labor force population aged >=16 y unemployed (unemployment rate), Percentage of families below the poverty level, Percentage of population below 138% of the poverty threshold, Percentage of single, Percentage of occupied housing units without a motor vehicle, Percentage of occupied housing units without a telephone, Percentage of occupied housing units without complete plumbing (log), Percentage of persons living in poverty, Unemployment rate, Per capita income, Percentage of persons at least 25 years old without a high-school diploma, Residential history derived - Opportunity Atlas Mean outcome for all children, Employment rate, Commute duration, Poverty rate, Public assistance rate, Homeownership rate, High-skill employment, Median household income, and Single-headed households.
- **Urbanization,** including Housing units per acre from EPA's Smart Location Database, Population Count Adjusted to Match 2015 Revision of UN WPP Country Totals in persons per 1 km2, Categorical measure of whether a census block is "urban" (2500 or more people) or "rural" (less than 2500 people), Composite index ranking census block groups according to their walkability, Traffic counts modeled at the 1 km2 resolution, Number of meters away from major road or highway.

### eResult 1 Determination of the optimal number of clusters (i.e., SDoH patterns) in the primary analysis.

In the primary analysis, we used multiple approaches to determine the optimal number of clusters, i.e., SDoH patterns, underlying the multidimensional, heterogenous SDoH data. First, the 'NbClust' suggested 3 potential clusters within the data, according to majority voting based on the 14 cluster measurements. To avoid a cluster, i.e., SDoH pattern dominating in the studied population, we split the large cluster (containing over 64% children in the study cohort) into 2 clusters, then we got 4 clusters (SDoH patterns). Importantly, we also observed clear 4-cluster structure of our data based on the dendrogram (**eFigure 1**) produced by the hierarchical clustering algorithm and the 2D t-SNE plot (**eFigure 2**). In conclusion, the optimal cluster (SDoH pattern) number was 4 in the primary analysis.

### eResult 2 SDoH pattern stability

In sensitivity analysis to dropped samples due to missing values, the hierarchical clustering algorithm detected the 4-cluster structure (**eFigures 3A and B**), very similar to that identified in the primary analysis. The re-identified clusters (i.e., SDoH patterns) were highly overlapped with that identified in the primary analysis (**eFigure 3C**). In addition, the chord diagram demonstrated that the re-identified SDoH patterns have the same SDoH profiles as that identified in the primary analysis (**eFigure 3D**).

In sensitivity analysis to include data samples. We first split the entire cohort into 5 folds, then each time we successive dropped 1 fold and used the remaining 4 folds to construct a subset to re-identify SDoH patterns. This is similar to the cross-validation strategy. In each subset, the cluster was re-identified using hierarchical clustering and the cluster number was determined using the same criteria in the primary analysis. We identified 4 SDoH patterns in each subset, which were also highly overlapped with the SDoH patterns identified in the primary analysis (**eFigures 4B and C**).

**eFigure 1 Dendrogram for SDoH pattern identification in the primary analysis.**

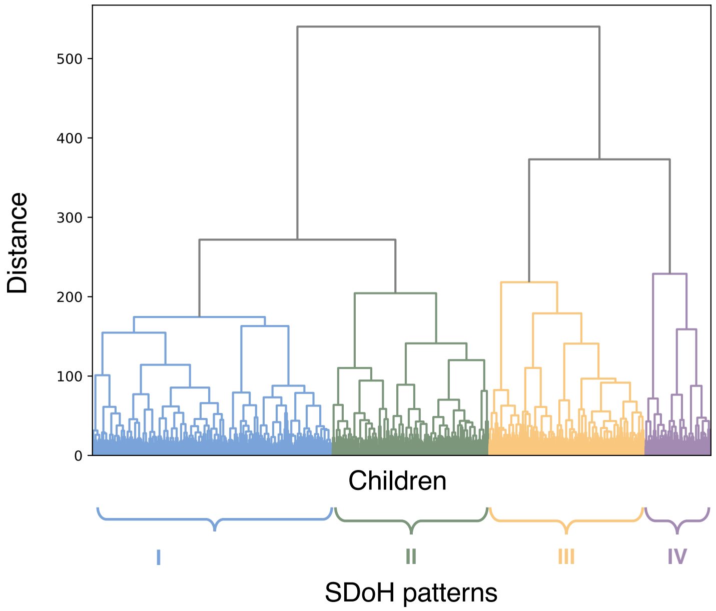

**eFigure 2 Visualization of SDoH patterns in 2D t-SNE space.**

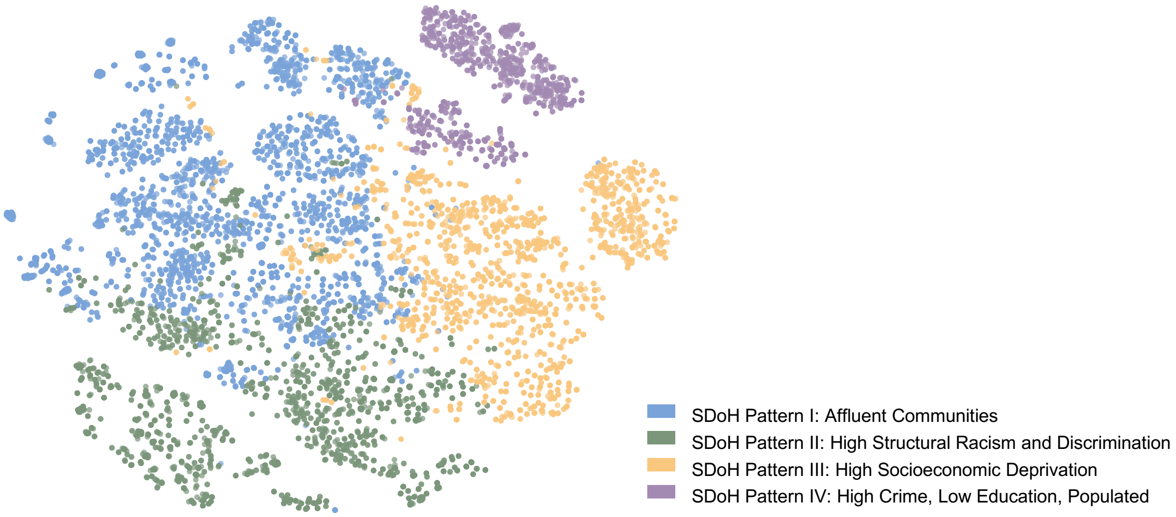

**eFigure 3 Results of sensitivity analysis to dropped samples due to missing values.**

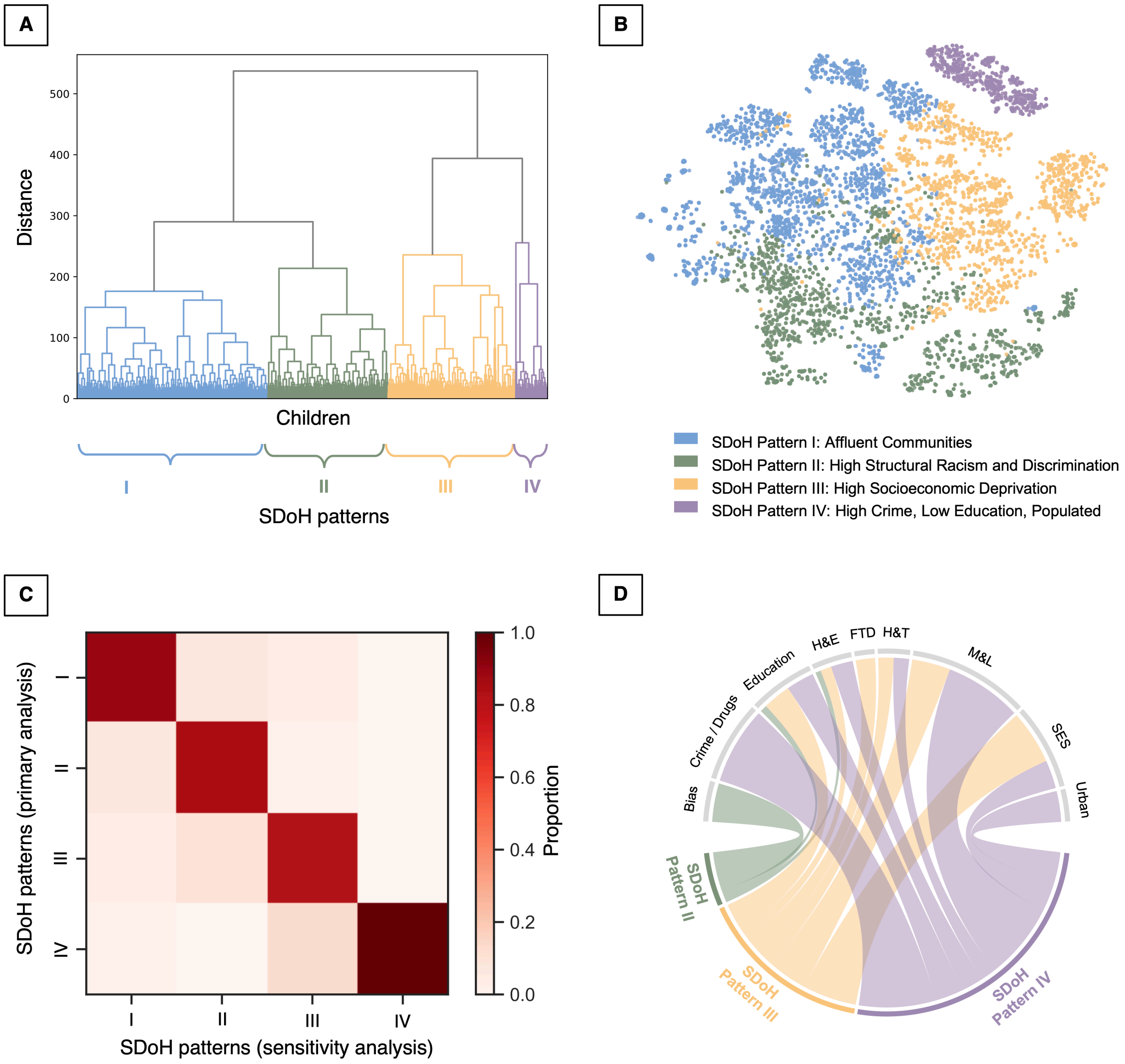

In this sensitivity analysis, we included children whose SDoH variables are with a missing rate < 70%. Same to the primary analysis, the SDoH variables were scaled based on z-score. K-nearest neighbors (KNN) imputation[23] was used to address missing values. After that, hierarchical clustering analysis was used to identify SDoH patterns. **A.** Dendrogram of hierarchical clustering analysis. **B.** Visualization of identified SDoH patterns in the 2D t-SNE space. **C.** Confusion matrix for comparing the SDoH patterns identified by the primary analysis and sensitivity analysis. Density of color indicates proportion of overlapped children within SDoH patterns identified by the primary analysis and sensitivity analysis. **D.** Chord diagram showing SDoH profiles of the re-identified SDoH patterns in the sensitivity analysis.

Abbreviations: FTD, Family type & Disability; H&E, Health & Environment; H&T, Housing type & Transportation; M&L, Minority status & Language; SES, socioeconomic status; SDoH, social determinants of health

**eFigure 4. Results of sensitivity analysis to included data samples.**

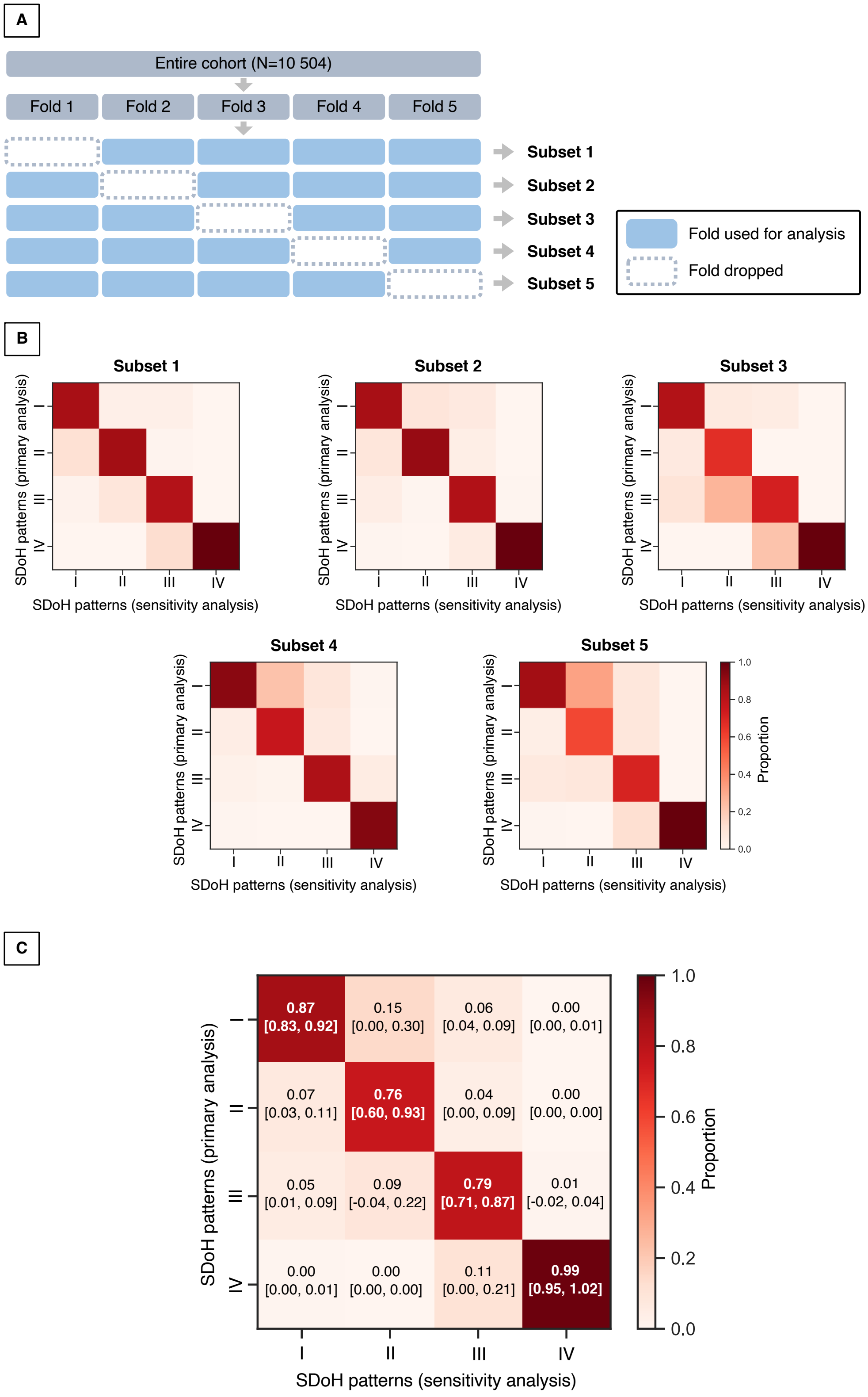

In this sensitivity analysis, (**A**) we first split the entire cohort into 5 folds, then each time we successive dropped 1 fold and used remaining 4 folds to construct a subset, which was used to re-identify SDoH patterns using hierarchical clustering analysis. **B.** Confusion matrices for comparing the SDoH patterns identified by the primary analysis and sensitivity analysis in each subset. Density of color indicates proportion of overlapped children within SDoH patterns identified by the primary analysis and sensitivity analysis. **C.** Averaged confusion matrix by aggregating results in the 5 subsets. Specifically, each color density of element in the confusion matrix is the mean value of corresponding elements across the 5 subsets. We also reported 95% confidence intervals.

### eTable 1 The 84 SDoH variables used for SDoH pattern identification.

| Variable | Missing rate | Median [IRQ] | Description |
|---|---|---|---|
| reshist_addr1_adi_edu_l | 0.05 | 2.49 [0.97-5.77] | Residential history derived - Area Deprivation Index: Percentage of population aged >=25 y with <9 y of education. |
| reshist_addr1_adi_edu_h | 0.05 | 92.27 [84.38-96.18] | Residential history derived - Area Deprivation Index: Percentage of population aged >=25 y with at least a high school diploma. |
| reshist_addr1_adi_work_c | 0.05 | 94.56 [91.57-96.71] | Residential history derived - Area Deprivation Index: Percentage of employed persons aged >=16 y in white collar occupations. |
| reshist_addr1_adi_income | 0.05 | 72367.00 [49786.00-97041.25] | Residential history derived - Area Deprivation Index: Median family income |
| reshist_addr1_adi_in_dis | 0.05 | 2.06 [1.26-2.96] | Residential history derived - Area Deprivation Index: Income disparity defined by Singh as the log of 100 x ratio of the number of households with <10000 annual income to the number of households with >50000 annual income. |
| reshist_addr1_adi_home_v | 0.06 | 221700.00 [145300.00-315100.00] | Residential history derived - Area Deprivation Index: Median home value. |
| reshist_addr1_adi_rent | 0.06 | 1042.00 [848.00-1314.00] | Residential history derived - Area Deprivation Index: Median gross rent. |
| reshist_addr1_adi_mortg | 0.06 | 1381.00 [1049.00-1716.00] | Residential history derived - Area Deprivation Index: Median monthly mortgage. |
| reshist_addr1_adi_home_o | 0.05 | 69.96 [50.17-83.50] | Residential history derived - Area Deprivation Index: Percentage of owner. |
| reshist_addr1_adi_crowd | 0.05 | 1.65 [0.50-3.83] | Residential history derived - Area Deprivation Index: Percentage of occupied housing units with >1 person per room (crowding). |
| reshist_addr1_adi_unemp | 0.05 | 7.50 [5.03-11.21] | Residential history derived - Area Deprivation Index: Percentage of civilian labor force population aged >=16 y unemployed (unemployment rate). |
| reshist_addr1_adi_pov | 0.05 | 7.15 [3.19-15.78] | Residential history derived - Area Deprivation Index: Percentage of families below the poverty level. |
| reshist_addr1_adi_b138 | 0.05 | 16.12 [9.06-30.20] | Residential history derived - Area Deprivation Index: Percentage of population below 138% of the poverty threshold. |
| reshist_addr1_adi_sp | 0.05 | 14.37 [8.92-23.46] | Residential history derived - Area Deprivation Index: Percentage of single. |
| reshist_addr1_adi_ncar | 0.05 | 4.96 [2.18-10.85] | Residential history derived - Area Deprivation Index: Percentage of occupied housing units without a motor vehicle. |
| reshist_addr1_adi_ntel | 0.05 | 1.65 [0.76-2.91] | Residential history derived - Area Deprivation Index: Percentage of occupied housing units without a telephone. |
| reshist_addr1_adi_nplumb | 0.05 | 0.00 [0.00-0.34] | Residential history derived - Area Deprivation Index: Percentage of occupied housing units without complete plumbing (log). |
| reshist_addr1_svi_pov_20142018 | 0.08 | 0.37 [0.15-0.68] | Residential history derived - Census tract CDC SVI (percentile % below poverty subcomponent) at primary residential address (ACS 2014-2018 5 yr avg) |
| reshist_addr1_svi_emp_20142018 | 0.08 | 0.41 [0.19-0.69] | Residential history derived - Census tract CDC SVI (percentile unemployment rate subcomponent) at primary residential address (ACS 2014-2018 5 yr avg) |
| reshist_addr1_svi_cap_20142018 | 0.08 | 0.38 [0.18-0.67] | Residential history derived - Census tract CDC SVI (percentile per capita income subcomponent) at primary residential address (ACS 2014-2018 5 yr avg) |
| reshist_addr1_svi_hs_20142018 | 0.08 | 0.34 [0.14-0.64] | Residential history derived - Census tract CDC SVI (percentile % no high school diploma subcomponent) at primary residential address (ACS 2014-2018 5 yr avg) |
| reshist_addr1_svi_65_20142018 | 0.08 | 0.39 [0.18-0.65] | Residential history derived - Census tract CDC SVI (percentile % persons 65 and older subcomponent) at primary residential address (ACS 2014-2018 5 yr avg) |
| reshist_addr1_svi_17_20142018 | 0.08 | 0.56 [0.30-0.81] | Residential history derived - Census tract CDC SVI (percentile % persons 17 and younger subcomponent) at primary residential address (ACS 2014-2018 5 yr avg) |
| reshist_addr1_svi_dis_20142018 | 0.08 | 0.33 [0.16-0.57] | Residential history derived - Census tract CDC SVI (percentile % population with a disability subcomponent) at primary residential address (ACS 2014-2018 5 yr avg) |
| reshist_addr1_svi_sin_20142018 | 0.08 | 0.47 [0.24-0.73] | Residential history derived - Census tract CDC SVI (percentile % single parent households with children under 18 subcomponent) at primary residential address (ACS 2014-2018 5 yr avg) |
| reshist_addr1_svi_min_20142018 | 0.08 | 0.49 [0.30-0.76] | Residential history derived - Census tract CDC SVI (percentile % minority subcomponent) at primary residential address (ACS 2014-2018 5 yr avg) |
| reshist_addr1_svi_eng_20142018 | 0.08 | 0.50 [0.28-0.74] | Residential history derived - Census tract CDC SVI (percentile % persons who speak English "less than well" subcomponent) at primary residential address (ACS 2014-2018 5 yr avg) |
| reshist_addr1_svi_hous20142018 | 0.08 | 0.59 [0.32-0.78] | Residential history derived - Census tract CDC SVI (percentile % housing structures with 10 or more units subcomponent) at primary residential address (ACS 2014-2018 5 yr avg) |
| reshist_addr1_svi_mob_20142018 | 0.08 | 0.00 [0.00-0.57] | Residential history derived - Census tract CDC SVI (percentile % mobile homes subcomponent) at primary residential address (ACS 2014-2018 5 yr avg) |
| reshist_addr1_svi_crwd20142018 | 0.08 | 0.47 [0.24-0.72] | Residential history derived - Census tract CDC SVI (percentile % crowding subcomponent) at primary residential address (ACS 2014-2018 5 yr avg) |
| reshist_addr1_svi_veh_20142018 | 0.08 | 0.44 [0.19-0.72] | Residential history derived - Census tract CDC SVI (percentile % households with no vehicle) at primary residential address (ACS 2014-2018 5 yr avg) |
| reshist_addr1_svi_grp_20142018 | 0.08 | 0.44 [0.00-0.68] | Residential history derived - Census tract CDC SVI (percentile % persons in group quarters) at primary residential address (ACS 2014-2018 5 yr avg) |
| reshist_addr1_opat_kfrpp_avg | 0.08 | 0.54 [0.46-0.60] | Residential history derived - Opportunity Atlas Mean outcome for all children |
| reshist_addr1_coi_zed_prxece | 0.08 | 0.64 [0.10-1.01] | Educational Domain High-quality early childhood education centers: Number of ECE centers within a 5-mile radius, converted to natural log units, transformed to z-scores. |
| reshist_addr1_coi_zed_prxhqece | 0.08 | 0.77 [0.58-0.88] | Educational Domain Access to healthy food: Number of NAEYC accredited centers within a 5-mile radius, converted to natural log units, transformed to z-scores. |
| reshist_addr1_coi_zed_ecenrol | 0.09 | 0.22 [-0.48-0.89] | Educational Domain High school graduation rate: Percentage 3- and 4-year-olds enrolled in nursery school, preschool or kindergarten, transformed to z-scores. |
| reshist_addr1_coi_zed_reading | 0.09 | 0.38 [-0.99-1.40] | Educational Domain School poverty: Percentage third graders scoring proficient on standardized reading tests, converted to NAEP scale score points, transformed to z-scores. |
| reshist_addr1_coi_zed_math | 0.09 | 0.32 [-1.30-1.55] | Educational Domain Third grade reading proficiency: Percentage third graders scoring proficient on standardized math tests, converted to NAEP scale score points, transformed to z-scores. |
| reshist_addr1_coi_zed_hsgrad | 0.09 | 0.36 [-0.51-0.86] | Educational Domain Third grade math proficiency: Percentage ninth graders graduating from high school on time, transformed to z-scores. |
| reshist_addr1_coi_zed_apenr | 0.08 | 0.42 [-0.14-1.19] | Educational Domain Adult educational attainment: Ratio of students enrolled in at least one AP course to the number of 11th and 12th graders, transformed to z-scores. |

| | | | |
|---|---|---|---|
| reshist_addr1_coi_zed_college | 0.09 | 0.42 [-0.16-1.01] | Educational Domain Early childhood education enrollment: Percentage 18-24 year-olds enrolled in college within 25-mile radius, transformed to z-scores. |
| reshist_addr1_coi_zed_schpov | 0.08 | 0.21 [-0.77-1.10] | Educational Domain Teacher experience: Percentage students in elementary schools eligible for free or reduced-price lunches, transformed to z-scores and multiplied by -1. |
| reshist_addr1_coi_zed_teachxp | 0.08 | -0.15 [-0.91-0.40] | Educational Domain Early childhood education centers: Percentage teachers in their first and second year, transformed to z-scores and multiplied by -1. |
| reshist_addr1_coi_zed_attain | 0.08 | 0.48 [-0.37-1.50] | Educational Domain College enrollment in nearby institutions: Percentage adults ages 25 and over with a college degree or higher, transformed to z-scores. |
| reshist_addr1_coi_zhe_food | 0.08 | 0.37 [-0.20-0.68] | Health and Environment Domain Access to green space: Percentage households without a car located further than a half-mile from the nearest supermarket, transformed to z-scores and multiplied by -1. |
| reshist_addr1_coi_zhe_green | 0.08 | -0.33 [-0.95-0.55] | Health and Environment Domain Extreme heat exposure: Percentage impenetrable surface areas such as rooftops, roads or parking lots, transformed to z-scores and multiplied by -1. |
| reshist_addr1_coi_zhe_walk | 0.08 | 0.47 [-0.43-1.37] | Health and Environment Domain Hazardous waste dump sites: EPA Walkability Index, transformed to z-scores. |
| reshist_addr1_coi_zhe_vacancy | 0.08 | 0.52 [-0.08-0.95] | Health and Environment Domain Walkability: Percentage housing units that are vacant, transformed to z-scores and multiplied by -1. |
| reshist_addr1_coi_zhe_suprfnd | 0.08 | 0.27 [0.27-0.27] | Health and Environment Domain Industrial pollutants in air, water or soil: Average number of Superfund sites within a 2-mile radius, converted to natural log units, transformed to z-scores and multiplied by -1. |
| reshist_addr1_coi_zhe_rsei | 0.08 | -0.40 [-0.71-0.49] | Industrial pollutants in air, water or soil: Index of toxic chemicals released by industrial facilities, converted to natural log units, transformed to z-scores and multiplied by -1. |
| reshist_addr1_coi_zhe_pm25 | 0.08 | 0.51 [-0.43-0.99] | Health and Environment Domain Housing vacancy rate: Mean estimated microparticle (PM2.5) concentration, transformed to z-scores and multiplied by -1. |
| reshist_addr1_coi_zhe_ozone | 0.08 | 0.64 [0.05-1.06] | Health and Environment Domain Airborne microparticles: Mean estimated 8-hour average ozone concentration, transformed to z-scores and multiplied by -1. |
| reshist_addr1_coi_zhe_heat | 0.08 | 0.71 [0.27-0.93] | Health and Environment Domain Health insurance coverage: Summer days with maximum temperature above 90F, transformed to z-scores and multiplied by -1. |
| reshist_addr1_coi_zhe_hlthins | 0.08 | 0.86 [0.29-1.21] | Health and Environment Domain Ozone concentration: Percentage individuals ages 0-64 with health insurance coverage, transformed to z-scores. |
| reshist_addr1_coi_zse_emprat | 0.08 | 0.57 [-0.00-1.03] | Employment rate: Percentage adults ages 25-54 who are employed, transformed to z-scores. |
| reshist_addr1_coi_zse_jobprox | 0.08 | 0.42 [-0.18-0.78] | Commute duration: Percentage workers commuting more than one hour one way, transformed to z-scores and multiplied by -1. |
| reshist_addr1_coi_zse_povrate | 0.08 | 0.48 [-0.26-0.89] | Social and Economic Domain: Poverty rate z-score |
| reshist_addr1_coi_zse_public | 0.08 | 0.42 [-0.43-0.84] | Social and Economic Domain: Public assistance rate z-score |
| reshist_addr1_coi_zse_home | 0.08 | 0.07 [-0.85-0.73] | Social and Economic Domain: Homeownership rate z-score |
| reshist_addr1_coi_zse_occ | 0.08 | 0.50 [-0.35-1.37] | Social and Economic Domain: High-skill employment z-score |
| reshist_addr1_coi_zse_mhe | 0.08 | 0.08 [-0.53-0.91] | Social and Economic Domain: Median household income z-score |
| reshist_addr1_coi_zse_single | 0.08 | 0.27 [-0.55-0.90] | Single-headed households: Percentage family households that are single-parent headed, transformed to z-scores and multiplied by -1. |
| reshist_addr1_p1tot | 0.05 | 5240.33 [1460.00-11593.33] | Residential history derived - Uniform Crime Reports: total adult offenses 1 |
| reshist_addr1_p1vlnt | 0.05 | 1084.00 [200.67-1947.67] | Residential history derived - Uniform Crime Reports: adult violent crimes 1 |
| reshist_addr1_drugtot | 0.05 | 2693.67 [840.67-6573.67] | Residential history derived - Uniform Crime Reports: durg abose violations total 1 |
| reshist_addr1_drgsale | 0.05 | 493.00 [106.00-1095.67] | Residential history derived - Uniform Crime Reports: drug sale total 1 |
| reshist_addr1_mjsale | 0.05 | 118.00 [50.33-437.33] | Residential history derived - Uniform Crime Reports: Marijuana sale 1 |
| reshist_addr1_drgposs | 0.05 | 1993.00 [590.33-5314.67] | Residential history derived - Uniform Crime Reports: drug possession total 1 |
| reshist_addr1_dui | 0.05 | 2265.00 [751.33-4499.00] | Residential history derived - Uniform Crime Reports: DUI 1 |
| reshist_addr1_leadrisk | 0.05 | 5.00 [2.00-8.00] | Estimated lead risk in census tract of primary residential address (1-10 scale) |
| reshist_state_sexism_factor | 0.00 | -0.37 [-0.93-0.61] | State level indicators of sexism from survey and implicit bias measures |
| reshist_state_racism_factor | 0.00 | -0.31 [-0.44-0.49] | State level indicators of racism from survey and implicit bias measures and state level structural variables |
| reshist_state_so_factor | 0.00 | -0.16 [-1.17-0.32] | State level indicators of bias against sexual orientation from structural variables |
| reshist_state_immigrant_factor | 0.00 | -0.45 [-0.62--0.16] | State level indicators of immigrant bias from survey and implicit bias measures and state level structural variables |
| reshist_state_mj_laws | 0.00 | 2.00 [2.00-3.00] | Marijuana state law during the same year as the assessment |
| reshist_addr1_d1a | 0.05 | 2.64 [1.00-4.85] | Residential history derived - gross residential density 1 |
| reshist_addr1_popdensity | 0.05 | 1666.72 [803.61-2805.40] | Residential history derived - UN adjusted population density 1 |
| reshist_addr1_urban_area | 0.05 | 1.00 [1.00-1.00] | Census Tract Urban Classification at current address #1 |
| reshist_addr1_walkindex | 0.05 | 10.67 [7.17-14.17] | Residential history derived - national walkability index 1 |
| reshist_addr1_traffic_count | 0.05 | 9546.69 [5404.09-15977.38] | Average Annual Daily Traffic Counts at current address #1 |
| reshist_addr1_proxrd | 0.05 | 826.30 [384.02-1556.07] | Residential history derived - proximity to major roads, in meters 1 |
| reshist_addr1_pm25_2016daysepa | 0.08 | 0.00 [0.00-1.00] | Residential history derived - number of days in 2016 PM2.5 was above EPA daily standards (35) at primary residential address at 1x1km2 |
| reshist_addr1_no2_2016_aavg | 0.05 | 18.82 [14.76-22.20] | Residential history derived - annual average of NO2 in 2016 at primary residential address at 1x1km2 |
| reshist_addr1_o3_2016_annavg | 0.05 | 40.59 [38.35-45.45] | Residential history derived - annual average of O3 in 2016 at primary residential address at 1x1km2 |

**eTable 2. Details of fitted models for mental health trajectory analysis by SDoH patterns (table can be found in the isolated Excel file).**

For outcomes in CBCL mental health symptom scores (continuous value), linear mixed effect models were used. For suicidal behavior outcomes (binary value), mixed effect logistic models were used. Individual and study sites were considered as two-level random effects. Sex and race/ethnicity were considered covariates. For race/ethnicity, white was considered as the reference group.
